# From Chest Pain to Coronary Functional Testing: Clinical and Economic Impact of Coronary Microvascular Dysfunction

**DOI:** 10.1101/2024.01.09.24301078

**Authors:** Ilan Merdler, Ryan Wallace, Andrew Hill, Giorgio A. Medranda, Pavan Reddy, Cheng Zhang, Sevket Tolga Ozturk, Vaishnavi Sawant, William S. Weintraub, Kassandra Lopez, Itsik Ben-Dor, Ron Waksman, Hayder D. Hashim, Brian C. Case

## Abstract

**Background:** Coronary functional testing to formally diagnose coronary microvascular dysfunction (CMD) reduces cardiovascular events and alleviates angina. This study aims to investigate the extensive and complex journey that patients with CMD undergo, from the onset of chest pain to eventual diagnosis.

**Methods:** Data from the Coronary Microvascular Disease Registry (CMDR) were analyzed, including information on the date of first documentation of chest pain, number of non-invasive and invasive tests the patient underwent, emergency department visits, and hospitalizations. In addition, we estimated the total cost per patient. A total of 61 patients with CMD diagnosis were included in this analysis.

**Results:** The cohort had an average age of 65.6±9.9 years. The median time from initial chest pain symptoms to diagnosis was 0.62 (interquartile range [IQR]: 0.06-2.96) years. During this period, patients visited the emergency department a median of 1.0 (IQR: 0.0-2.0) times. Diagnostic tests included 3.0 (IQR: 2.0-6.0) electrocardiograms, 3.0 (IQR: 0.0-6.0) high-sensitivity troponin tests, and 1.0 (IQR: 1.0-2.0) echocardiograms. Prior to diagnosis of CMD, 13 (21.3%) patients had left heart catheterization without coronary functional testing. Non-invasive testing for ischemia was conducted in 43 (70.5%) patients. Alternative non-cardiac diagnoses were given to 11 (18.0%) patients during the diagnostic process, with referrals made to gastroenterology for 16 (26.2%) and pulmonology for 10 (16.4%) patients. The cost averaged $1,790±2,506 per patient.

**Conclusion:** Timely identification of CMD offers promising opportunities for prompt symptom alleviation, accompanied by reduced visits to the emergency department, cardiovascular testing, invasive medical procedures, and consequently reduced healthcare expenses.

**Clinical Trial Registry:** Coronary Microvascular Disease Registry (CMDR), clinicaltrials.gov, NCT05960474

**What is Known; What the Study Adds:** *What is Known:*

- Angina pectoris is a major global health concern, impacting millions of individuals around the world.
- Coronary microvascular dysfunction (CMD) is a known etiology to cause angina.

*What the Study Adds:*

- This study reveals the challenging journey of CMD patients from chest pain to diagnosis, showing the complexity, and overlapping symptoms of CMD, leading to under/misdiagnosis or delay in definitive diagnosis.
- Healthcare providers must improve CMD awareness and understanding to ensure timely and accurate diagnosis, minimizing patient burden and unnecessary expenses.
- Further research and awareness campaigns are crucial to optimize CMD management, leading to better healthcare outcomes and reduced economic strain.

## Introduction

Angina pectoris is a major global health concern, impacting millions of individuals around the world^1^. It is a prevalent symptom of myocardial ischemia, highlighting the need for prompt, accurate diagnosis. While the conventional approach to diagnosing this condition involves coronary angiography to detect coronary artery stenosis, there is a tendency to overlook non-obstructive coronary artery disease and disregard the potential presence of coronary microvascular dysfunction (CMD)^2, 3^. This oversight presents an ongoing challenge in clinical practice, hampering effective management and treatment strategies.

Despite the availability of appropriate devices and techniques for coronary functional testing to diagnose CMD, there is a lack of utilization and insufficient prioritization of these assessments in evaluating patients with angina pectoris^4^. Additionally; existing guidelines primarily focus on non-invasive and invasive diagnostic approaches for patients with stable angina evaluating for epicardial stenosis, neglecting the crucial aspect of when to perform coronary functional testing to evaluate the coronary microvasculature^5^. The broader adoption of coronary functional tests has various obstacles, including limited awareness, overreliance on established diagnoses, technical complexities, and potential risks associated with invasive procedures^6–8^. Consequently, overlooked patients frequently end up with repeated visits to the emergency department, receiving multiple cardiovascular tests, and redundant invasive procedures, raising healthcare costs^9, 10^. Comprehensive assessment of both the epicardial vessels and microvasculature (coronary functional testing) has the potential to reduce cardiovascular events, alleviate angina symptoms, decrease hospitalizations, decrease cardiovascular testing, and reduce healthcare costs.

To tackle these challenges, the Coronary Microvascular Disease Registry (CMDR) has emerged as an innovative initiative. This prospective, multi-center, and standardized registry aims to enroll patients with angina and non-obstructive coronary artery disease who undergo invasive hemodynamic assessment of the coronary microvasculature. The current study aims to shed light on the complex and lengthy journey that patients with CMD experience, from the onset of chest pain to the eventual diagnosis (Figure 1).

**Figure 1.**
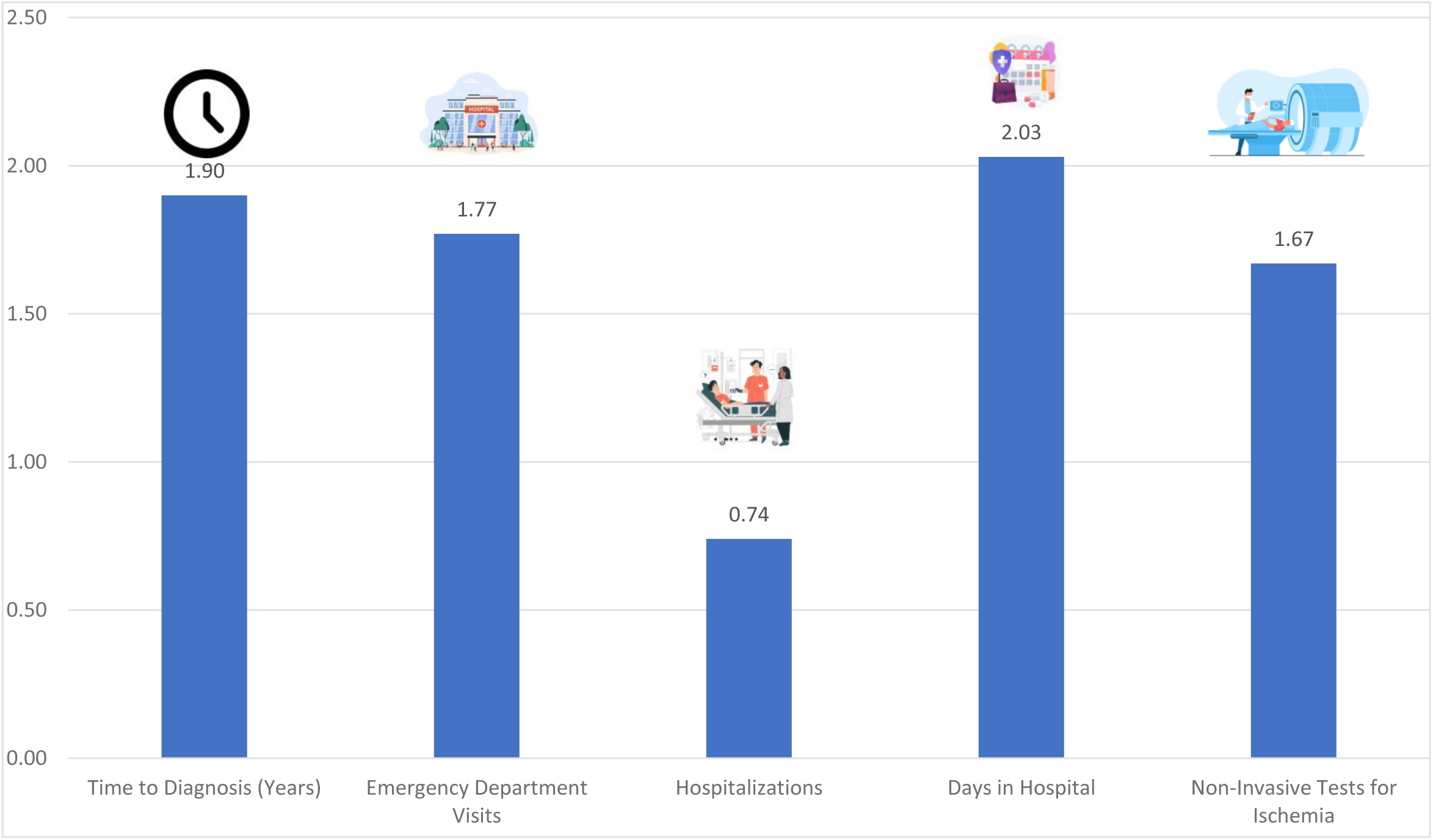
The journey an average patient with coronary microvascular dysfunction undergoes. Illustration depicting the diagnostic journey of patients experiencing chest pain until the diagnosis of coronary microvascular dysfunction (CMD) is reached, spanning nearly two years. The figure highlights the average days till diagnosis, emergency room visits, hospitalizations, and days in hospital during this period, showcasing the complexity and duration of the diagnostic process.

- Pictures were taken from freepik.com
- The numbers represent the average visits
- The number of non-invasive tests for ischemia is calculated by the total number of any non-invasive tests each patient divided by the number of patients

## Methods

The CMDR was established to comprehensively investigate patients experiencing chest pain and non-obstructive coronary artery disease who undergo invasive hemodynamic assessment of the coronary microvasculature, including patients with INOCA (ischemia with no obstructive coronary arteries) and MINOCA (myocardial infarction with no obstructive coronary arteries)^11^. Data for the registry were gathered from established hospital or clinical care databases. No research-related interventional procedures were performed solely for the purpose of this study; instead, the procedures were based on clinical judgment and tailored to each patient’s specific needs. The registry strictly adhered to the guidelines provided by the MedStar Health Institutional Review Board. The CMDR has been registered on ClinicalTrials.gov under the identifier NCT05960474.

Patient enrollment took place at two MedStar Health centers, MedStar Washington Hospital Center in Washington, DC, and MedStar Southern Maryland Hospital Center in Clinton, MD. The first patient was enrolled in the registry in August 2021, and data from through November 2023 were included in the analysis.

To assess coronary microvasculature physiology, we used the Coroventis CoroFlow Cardiovascular System (Abbott Laboratories, Chicago, Illinois). Non-hyperemic resting indices, specifically the resting full-cycle ratio, and hyperemic index, fractional flow reserve, were measured using a physiologic pressure wire (PressureWire™ X Guidewire, Abbott Laboratories). Additionally, we utilized thermodilution technology to evaluate coronary flow reserve (CFR), the index of microvascular resistance (IMR), and the resistive reserve ratio^12^. All measurements were recorded and incorporated into the CMDR alongside the corresponding findings from coronary angiography.

Patients diagnosed with CMD were identified based on specific criteria: a CFR value <2.5 and an IMR ≥25^13^. To ensure accurate classification, cases falling within the borderline range were evaluated by a multidisciplinary heart team, which determined the likelihood of CMD^14^.

We collected comprehensive data regarding baseline characteristics, co-morbidities, medications, severity of chest pain, non-invasive cardiovascular tests, coronary anatomy based on angiography, physiologic measurements, and post-procedural outcomes. Furthermore, detailed information regarding medications used during catheterization, procedure time, and the amount of contrast and radiation were documented. Data were also collected regarding the date of first documentation of chest pain, non-invasive and invasive testing performed for cardiac and non-cardiac reasons, emergency department visits, and hospital stays. The primary outcome was time from chest pain until diagnosis, and secondary outcomes were hospital visits and tests performed during the period leading up to diagnosis. Lastly, we estimated cost based on the available online payment information (for Medicare, using cms.gov) pertaining to various medical procedures and services^15^. These include the cost of an emergency department visit, hospital stay, electrocardiogram (EKG), troponin laboratory test, nuclear stress test, transthoracic echocardiogram, cardiac magnetic resonance imaging (MRI), computed tomography (CT) calcium score, coronary CT, stress treadmill EKG, stress echocardiography, left heart catheterization, and office visits. It is crucial to acknowledge that real-life costs are multifaceted, encompassing insurance companies, providers, operators, and patients. In particular, Medicare payments do not include professional fees. Obtaining an accurate estimate of costs can be challenging, and our analysis, though simplistic and non-specific, was intended solely to offer a general understanding of potential additional expenses resulting from delays in diagnosis.

The results are reported as sample mean and sample standard deviation and sample median and interquartile range for continuous variables and as a percentage for binary variables.

## Results

The current analysis consists of a comprehensive evaluation of 234 patients enrolled in the CMDR. Due to inadequate documentation regarding the process between chest pain and diagnosis, three CMD-positive patients were excluded from the current study, yielding 61 patients identified as CMD-positive. Table 1 provides a concise summary of the baseline characteristics of these participants. Most patients were female (46/61, 75.4%) or African American (33/61, 54.1%), and the cohort had a high prevalence of associated cardiovascular co-morbidities. The patients’ mean age was 65.6±9.9 years.

**Table 1.** Baseline Characteristics.

| **Variable** | **CMD positive (N=61)** |
| --- | --- |
| Age (years) | 65.6±9.9 |
| Body Mass Index (kg/m<sup>2</sup>) | 28.0±5.8 |
| Female | 46/61 (75.4%) |
| Caucasian | 23/61 (37.7%) |
| African American | 33/61 (54.1%) |
| Hispanic | 2/61 (3.3%) |
| Tobacco use (current and past) | 21/60 (35.0%) |
| Hypertension | 52/61 (85.2%) |
| Hyperlipidemia | 45/61 (73.8%) |
| Diabetes | 15/61 (24.6%) |
| Chronic Kidney Disease (GFR<60mL/min/1.73m<sup>2</sup>) | 6/61 (9.8%) |
| Peripheral Arterial Disease | 5/61 (8.2%) |

Values are n (%) or mean ± SD
GFR=glomerular filtration rate

The median time from initial chest pain symptoms to diagnosis was 0.62 (interquartile range [IQR]: 0.06-2.96) years. A detailed account of the events that occurred from the onset of chest pain until diagnosis is provided in Table 2. During the period leading up to the diagnosis, patients visited the emergency department a median of 1.0 (IQR: 0.0-2.0) times.

**Table 2.** Diagnostic procedures till CMD assessment.

| Variable | CMD positive (N=61) |  |
| --- | --- | --- |
| | Mean ± SD | Median [IQR] |
| Time (years) | 1.9±2.7 | 0.62 [0.06-2.96] |
| ED visits | 1.8±2.7 | 1.0 [0.0-2.0] |
| Chest pain with hospital stay (events) | 0.7±1.3 | 0.0 [0.0-1.0] |
| Hospital-stay (days) | 2.0±4.4 | 0 [0.0-2.0] |
| EKG | 6.1±7.9 | 3.0 [2.0-6.0] |
| Blood analysis (any) | 9.3±15.1 | 5.0 [2.0-11.0] |
| Echocardiogram | 1.5±1.8 | 1.0 [1.0-2.0] |
| LHC | 13/61 (21.3%) | - |
| NST | 35/61 (57.4%) | - |
| Cardiac MRI | 4/61 (6.5%) | - |
| Calcium Score CT | 14/61 (23.0%) | - |
| Coronary CT | 16/61 (26.2%) | - |
| CT – PE protocol | 15/61 (24.6%) | - |
| Non-cardiac diagnosis | 11/61 (18.0%) | - |

\*ED=Emergency Department; EKG=Electrocardiogram; LHC=Left Heart Catheterization; NST=Nuclear stress test; MRI=Magnetic resonance imaging; CT=Computed tomography; PE=Pulmonary embolism.
\*\* Hospital stay in days was calculated according to total days in hospital including repeat visits
\*\*\*Median numbers hold greater importance in cases of abnormal distribution; however, we also provide mean and standard deviation (SD) for the convenience of the readers.

Throughout this extended diagnostic journey, patients underwent multiple diagnostic tests and procedures. Specifically, the median number of EKG tests was 3, median number of blood analyses (of any kind) was 5, median number of echocardiograms was 1,and median number of high-sensitivity troponin tests was 3. Further, 13 out of 61 patients (21.3%) underwent invasive left heart catheterization without coronary functional testing.

Among the patient cohort, 43 (70.5%) underwent non-invasive testing for ischemia. Notably, the frequency of testing varied, with some patients receiving multiple tests, while others did not undergo any such evaluations. On average, each patient had 1.7±2.3 non-invasive tests for ischemia. Nuclear stress tests were performed on 35 patients (57.4%), with 12 out of 35 (34.3%) yielding positive results. Cardiac MRI was conducted on 4 patients (6.5%), of which 1 out of 4 (25%) showed positive findings. Additionally, coronary CT was performed on 16 patients (26.2%), with 3 out of 16 (18.8%) demonstrating a significant lesion. Calcium score CT scans were conducted on 14 patients (23.0%), and 5 out of 14 (35.7%) showed a high calcium burden, with an average calcium score of 160±162 Agatston units. CT in the pulmonary embolism protocol was performed on 15 patients (24.6%). Among the study participants, only two patients were referred to an electrocardiogram treadmill stress test, two to cardiopulmonary exercise testing, and 7 out of 61 (11.5%) were referred to exercise stress echocardiography. None of the patients underwent dobutamine stress echocardiography.

In total, 9 out of 61 patients (18.0%) received an alternate non-cardiac diagnosis during the diagnostic process. Among the cohort, 16 out of 61 patients (26.2%) were referred to gastroenterologist (GI) experts and subsequently underwent endoscopic assessment. Furthermore, 10 out of 61 patients (16.4%) were referred to pulmonology specialists for further evaluation.

The sample average cost per patient was $1,790±2,506. This estimate is derived from presumed costs based on the Medicare fee-for-service schedule and is intended as a generalized assumption, not encompassing the entire economic burden associated with chest pain on different healthcare systems. Table S1 provides a summary of our calculation method, considering various factors such as ED visits, hospital stays, EKGs, troponin laboratory tests, stress tests, echocardiograms, imaging, left heart catheterization, and non-cardiac office visits (GI and pulmonary).

## Discussion

Our study investigating the journey of CMD patients from initial symptoms to final diagnosis yielded several notable findings. First, we observed that the median time for CMD to be formally diagnosed is approximately six months, and in that time period the majority of patients had numerous hospital visits and hospitalizations. This prolonged duration highlights the challenges of accurately identifying and diagnosing CMD, and highlighting that providers need to keep CMD in mind as a possible diagnosis. Second, we noted a significant number of non-invasive tests conducted during this diagnostic period. These tests were employed to understand and assess the patients’ condition but without arriving at the formal diagnosis. Third, it was concerning to discover that a quarter of patients underwent left heart catheterization at first without undergoing coronary functional testing. This omission may have resulted in missed opportunities for a comprehensive evaluation, despite exposing the patient to increased risk with an invasive procedure, and potentially led to delayed or inaccurate diagnoses. Furthermore, our study showed that the costs of delayed diagnosis might add up to thousands of dollars.

It is crucial for physicians to conduct a proper differential diagnosis for patients experiencing chest pain^16^. If coronary artery disease is suspected, based on symptoms, the next step is to determine whether invasive or non-invasive assessment is necessary^17^. Patients with chest pain should be suspected of having CMD early on, even before knowledge about their coronaries. Further, vasospastic angina should also be considered^18^. Based on previous trials and the current study, it has been observed that certain patients with specific characteristics, such as being female and having cardiovascular risk factors, are more commonly associated with CMD. The reasons for this gender disparity are not entirely clear, but hormonal factors, differences in blood vessel structure, and other physiological factors may play a role^19^. Patients with established cardiovascular risk factors, such as hypertension, diabetes, dyslipidemia, and obesity, are at a higher risk of developing CMD. These risk factors can contribute to endothelial dysfunction and inflammation in the small blood vessels of the heart, further impairing their ability to function correctly^20^. Overall, by recognizing that CMD is more commonly seen in females and those with CV risk factors, healthcare providers can be vigilant in considering this differential diagnosis and providing appropriate care for affected patients. Early detection and intervention are critical to improving outcomes and quality of life for individuals with CMD.

Addressing the impact on the quality of life is crucial in the context of limiting chest pain^21^. Unfortunately, patients may persistently experience chest pain without receiving appropriate therapy while undergoing various, repeated tests. In some instances, they might even be prescribed medications and treatments that are unsuitable and carry potential harm. The absence of clear guidelines on optimal timing to evaluate for CMD prolongs this distressing situation for patients, potentially stretching it out over many months to even a year. Consequently, these individuals may gradually develop a sense of mistrust toward the healthcare system as their symptoms persist without a definitive diagnosis^22^. The patient becomes frustrated with their provider as they cannot find etiology for their symptoms, resulting in the seeking a second opinion from another provider, usually coming to the same conclusion. This leads to patient’s having an overall mistrust to the healthcare system. Further, providers become frustrated when they are not able to provide the correct diagnosis, and subsequent treatment, for a patient suffering from a specific symptoms, such as angina. Knowledge about CMD, and making the subsequent diagnosis, will lead to decreasing this mistrust, and frustration, for both the patient and the provider.

The treatment of CMD is complex and primarily relies on medical therapy, such as beta blockers and calcium channel blockers. However, several novel experimental therapies are under investigation and may provide relief for these patients^23^. Proper identification, early diagnosis, and treatment of CMD could reduce hospital visits and improve quality of life. We have highlighted that there may be significant expenses incurred during the period until the true diagnosis is made. Further, a delay in diagnosis can also lead to repeat invasive procedures, which might put the patient at risk for complications or repeated CTs which could lead to unnecessary radiation exposure.

Coronary functional testing plays a crucial role in evaluating the complex nature of the microvasculature, encompassing not only CMD but also other components such as coronary vasospasm and structural and functional abnormalities. While CMD is an essential disease process of microvascular health, it is equally important to understand the broader spectrum of coronary functional abnormalities for comprehensive patient assessment. Therefore, in addition to measuring CFR and IMR, implementing a vasospastic challenge with acetylcholine should be considered to further evaluate and differentiate between vasospastic angina and CMD in cases of unexplained chest pain. This comprehensive approach ensures a more accurate and thorough assessment of the underlying condition, enabling clinicians to tailor treatment strategies accordingly^24^.

Collaborative efforts are crucial to advance the field of CMD research. The establishment of nationwide or international CMD registries, such as the CMDR used in our study, promotes the pooling of data from diverse patient populations and healthcare settings. The CMDR serves as a platform for patients undergoing testing for microvascular disease using devices approved for marketing by the US Food and Drug Administration. Data submission and analysis are free for all sites interested in participating, and a follow-up module will be added but will require patient consent. This registry enables a more comprehensive understanding of CMD, its clinical presentation, treatment patterns, and outcomes across different populations.

Several limitations should be acknowledged. First, the cohort size was small, and the study was primarily descriptive. Second, patient selection for CMD assessment was left to the operator’s discretion, introducing potential selection bias. The cost analysis conducted in this study has some limitations; specifically, we did not assess the cost-effectiveness of these evaluations, which could provide valuable insights into the overall efficiency of the procedures. Moreover, it is important to note that our estimate did not account for various other components that might significantly impact the total cost per patient. Healthcare economics is a complex field, and our analysis was primarily focused on specific medical procedures and readily available online information for the selected study population. As a result, there are potential factors beyond the scope of this paper that could influence the total costs. Therefore, it is vital to acknowledge the inherent limitations in our study and recognize that this estimate may not fully capture all possible cost-related factors associated with the diagnostic evaluation of CMD. Future research should strive to address these limitations and incorporate a broader range of factors for a more comprehensive and accurate cost assessment. However, despite these limitations, our study represents the foundation and future potential of a nationwide, or even international, CMD registry and helps establish a proper time limitation for checking other causes of chest pain before CMD assessment.

In conclusion, our study sheds light on the journey that patients with CMD endure from the onset of chest pain to diagnosis. Our findings emphasize the critical need for improved awareness and understanding of CMD among healthcare providers. Timely and accurate diagnosis is crucial to minimize the burden on patients, both in terms of their health outcomes and the financial implications of unnecessary procedures. Further research and awareness campaigns are necessary to enhance the diagnostic process and optimize the management of CMD patients, ultimately leading to improved overall healthcare outcomes and reduced economic strain.

## Data Availability

The data that support the findings of this study are available on request from the corresponding author.

## Acknowledgments

The authors acknowledge Jason Wermers, MS, a paid medical writer and employee of MedStar Health, for editing and preparing the manuscript for submission.

## Disclosures

### Sources of Funding

The Coronary Microvascular Disease Registry is an investigator-initiated study funded by MedStar Health Research Institute.

### Conflicts of Interest

Brian C. Case, reports being a speaker for Zoll Medical.

Hayder Hashim reports serving on the advisory boards of, and being a speaker for, Abbott Vascular, Boston Scientific, and Philips IGT.

Ron Waksman reports serving on the advisory boards of Abbott Vascular, Boston Scientific, Medtronic, Philips IGT, and Pi-Cardia Ltd.; being a consultant for Abbott Vascular, Biotronik, Boston Scientific, Cordis, Medtronic, Philips IGT, Pi-Cardia Ltd., Swiss Interventional Systems/SIS Medical AG, Transmural Systems Inc., and Venous MedTech; receiving institutional grant support from Amgen, Biotronik, Boston Scientific, Chiesi, Medtronic, and Philips IGT; and being an investor in MedAlliance and Transmural Systems Inc.

All other authors – None.

## Abbreviations

CFR: coronary flow reserve
CMD: coronary microvascular disease
CMDR: Coronary Microvascular Disease Registry
IMR: index of microvascular resistance

## Notes

### Clinical Trial

Coronary Microvascular Disease Registry (CMDR), clinicaltrials.gov, NCT05960474

### Author Declarations

The registry strictly adhered to the guidelines provided by the MedStar Health Institutional Review Board.

